# Empirical Approach to Developing an Optimal Socioeconomic Status Index for Health Surveillance

**DOI:** 10.1101/2022.01.14.22269310

**Authors:** Jordge LaFantasie, Francis Boscoe

## Abstract

The association between multi-dimensional deprivation and public health is well established, and many area-based indices have been developed to measure or account for socioeconomic status in health surveillance. The Yost Index, developed in 2001, has been adopted in the US for cancer surveillance and is based on the combination of two heavily weighted (household income, poverty) and five lightly weighted (rent, home value, employment, education and working class) indicator variables. Our objectives were to 1) update indicators and find a more parsimonious version of the Yost Index by examining potential models that included indicators with more balanced weights/influence and reduced redundancy and 2) test the statistical consistency of the factor upon which the Yost Index is based. Despite the usefulness of the Yost Index, a one-factor structure including all seven Yost indicator variables is not statistically reliable and should be replaced with a three-factor model to include the true variability of all seven indicator variables. To find a one-dimensional alternative, we conducted maximum likelihood exploratory factor analysis on a subset of all possible combinations of fourteen indicator variables to find well-fitted one-dimensional factor models and completed confirmatory factor analysis on the resulting models. One indicator combination (poverty, education, employment, public assistance) emerged as the most stable unidimensional model. This model is more robust to extremes in local cost of living conditions, is comprised of ACS variables that rarely require imputation by the end-user and is a more parsimonious solution than the Yost index with a true one-factor structure.

---

It has been long established in the literature that social determinants of health and multi-dimensional deprivation are strongly associated with health outcomes (Gilbert et al. 2003, Braveman et al. 2010, Phillips et al. 2016, Glassman 2019, Singu et al. 2020). Area based deprivation indices are a common method used to approximate affluence, particularly for the purpose of allocating funding or determining relationships in public health. They are especially valuable when individual level data are not available. The intent of these indices is to estimate relative deprivation for a given geography using a multidimensional approach to approximate social position and material access (Schuurman et al. 2007, Boscoe et al. 2021). The multidimensional nature ensures that the indices capture sources of deprivation beyond income and are less sensitive to measurement bias than univariate measures of affluence (Bryere et al. 2017, Glassman 2019). Indicators surrounding economic status (e.g., income, poverty), housing (home value, rent/mortgage, crowding, technology), social network (e.g., marital status, citizenship), employment, education and mobility are typically included in an index, although the number of indicators varies widely (Glassman 2021).

The Yost Index was introduced in 2001 (Yost et al. 2001) and has since been adopted in the US for cancer surveillance (Boscoe et al. 2021). It employs seven indicators: median household income, median house value, median gross rent, percent below 150% poverty, working class and percent unemployed, all of which are derived from US Census or American Community Survey data (National Cancer Institute 2021). It is most strongly influenced by measures of wealth (i.e. median household income, poverty, Table 1, Yu et al. 2014, Boscoe et al. 2021).

**Table 1.** Yost indicator variables and approximate beta weights derived from one-factor factor analysis according to Yu et al. (2014).

| Census Indicator | Beta Weight |
| --- | --- |
| % working class | -0.1 |
| % unemployed aged 16 and over | -0.05 |
| % of persons below 150% of poverty line | -0.25 |
| Median household income | 0.48 |
| Education index (weighted school years) | 0.1 |
| Median house value | 0.05 |
| Median rent | 0.06 |

The Yost Index was developed twenty years ago. Indices and their indicator variables can stale as conditions change; updates to indices should consider new indicators that may be more relevant considering evolving technologies and economics. For example, working class may be redundant to income and education, and the occupation classification system is susceptible to changes in career classification rules and diverse earning potentials within each class. Housing cost/value is temporally volatile, spatially autocorrelated, and may bias index values for areas with very high or very low cost of living. Instead, income disparity measures such as the Gini Index or housing cost burden (% of income dedicated to housing) might be indicative of disparities in socioeconomic status (SES) under various neighborhood and census tract economic and developmental conditions (Liao and De Maio 2021). Citizenship, nativity, and language proficiency, which are included in some deprivation indices (Guillaume et al. 2016), all influence access to resources and social networks and might be considered a source or covariate of affluence or deprivation. Access to plumbing and home telephone are components of some deprivation indices (Guillaume et al. 2016, Glassman 2019, Berg et al. 2021) and are likely relevant in many developing situations; these may be analogous to owning a laptop/desktop computer in the United States. Poor health is a form of deprivation; access to health insurance might be used as a proxy for health since health measures are not included in the ACS data (Glassman 2021). The need for public assistance/government benefits should also be considered (Deas et al. 2003, Phillips et al. 2016); application for public assistance is a nuanced process that might better inform an index than simple household income or poverty.

The weights applied to the indicators when calculating the index are just as important as the indicators themselves. If the weights are incorrectly calculated, the resulting index may not be measuring what it purports to measure. Weights for composite indices can be determined through a variety of techniques including “subjective/arbitrary” weighting based on expert opinion or survey or empirical weighting wherein analysts use multivariate regression or factor analysis to model a latent variable of interest (e.g. deprivation, SES) using various indicators/measured variables (Cowan et al. 2012, Mazziotta and Pareto 2013). The Yost Index uses indicator weights based on beta weights/factor loadings resulting from one-component principal components analysis or one-factor factor analysis (Yost et al. 2001, Yu et al. 2014). The fact that the Yost Index is dominated by monetary indicators (poverty, household income, Table 1) may indicate that the indicators included in the Yost index are not unidimensional and may require more than one factor to describe their overall variance. That is, a one-factor model may not be internally consistent/have a statistically reliable fit. This would not be unprecedented-recently, Berg et al. (2021) reported that the Area Deprivation Index, originally based on a unidimensional structure, did not fit one dimension. They revised ADI to the ADI-3 to include three factors rather than one and achieved better internal consistency and fit in the model.

## Objectives

Our objectives were to update indicators and find a more parsimonious version of the Yost Index by examining potential models that included indicators with more balanced weights/influence and reduced redundancy. A component of this objective was to determine the statistical reliability (internal consistency) of the Yost Index indicators as a one-factor model, and to find a parsimonious one-factor structure that matched the utility of the Yost Index.

## Methods

We used a multi-step process for model selection and verification:

1. Data processing and subsampling,
2. EFA and Indicator combination reducton
  a. Conduct exploratory one-factor factor analysis on all viable combinations of the fourteen potential indicator variables,
  b. Cull the possible indicator combinations to the best potential models using fit statistics, indicator weight balance and indicator inclusion,
3. Further model selection-CFA
  a. Conduct confirmatory factor analysis on the best models emerging from step 2,
  b. Further cull the potential models to one final model based on reliability, unidimensional nature and fit statistics,
4. Final Model Verification
  a. Further verify reliability of the final model via cross-validation with
    i. a subset (20%) test dataset,
    ii. a temporally distinct dataset
    iii. spatially distinct datasets
  b. Check for convergent validity by relating the final model index values to a related but distinct concept- affluence influenced health measures.
5. Upon verifying the internal reliability and external validity of the selected model we then compared the proposed model and the original Yost model and noted differences.

### Data access and processing

We gathered American Community Survey aggregated five-year data for the 2015-2019 timeframe at the census tract level for the United States (excluding territories) from NHGIS (Manson et al. 2020). We calculated Yost variables according to the SEER Yost index guidelines on their variables webpage https://seer.cancer.gov/seerstat/variables/countyattribs/time-dependent.html and additional potential indicators as detailed in Table 2.

**Table 2.** ACS table identification and calculation of potential indicators.

|  | Current/<br>Potential Indicator | Description | Table; Calculation |
| --- | --- | --- | --- |
| Current Yost Indicators | Education (edu) | Sex by education attainment for the population aged 25 years and older | Table B15002 |
|  | Poverty (pov) | Poverty status by sex by age | Table B17001 |
|  | Household income (mhi) | Household Income | Table B19001 |
|  | Employment (emp) | Employment status | Table B23025 |
|  | Rent (mgr) | Gross rent | Table B25063 |
|  | Home value (mhi) | Median owner-occupied home value | Table B25075 |
|  | Working Class (%) (wc) | Sex by occupation for the civilian employed population 16 years and older | Table C24010 |
| Potential Indicators | Gini Index (gini) | Measure of income disparity between 0 and 100. | The Gini index was calculated for most census tracts and provided in ACS data Table B19083 |
|  | Public Assistance (pa) | Receipt of public assistance by households (%) | Table B19057; With public assistance income/Total |
|  | Housing cost burden (housepct2) | Income allocated to housing (%; household) | Tables B25071, B25092; Average of Median gross rent as % of household income and Median selected monthly owner costs as % of household income total |
|  | Number of computers (comp) | Households without a laptop or desktop computer % | Table B28010; (No computer + no desktop or laptop)/ total |
|  | Language (lang) | Speak English “less than very well” (%) | Table C16001; Sum (All languages “less than very well”)/ total |
|  | Health insurance (hins) | No health insurance coverage (%) | Table B27020; (Native no ins+ naturalized no ins +noncitizen no ins)/total |
|  | Crowding (crowd) | More than two people per room (%) | Table B25014; (Owner + renter occupied 2.01 or more occupants per room)/ total |

We further pre-processed the ACS data according to Boscoe et al. (2021). Briefly, tracts with greater than 1/3 of the population represented by group quarters, tracts with populations less than 100 or with less than 30 housing units (1988), and tracts with more than three missings (2) were removed with the resulting number of tracts equaling 71066. Of the tracts remaining, 1964 (∼3%) had at least one missing value, a number of which were median home value, which, if removed in pair or row-wise calculations, would exclude census tracts comprised of public housing and primarily rentals. Thus, while 3% is a small number of missing values, we felt it critical to include all possible tracts by imputing missing data.

Missing values were imputed five times using the classification and regression tree (“cart”) method in the `mice` package in R (Van Buuren and Groothuis-Oudshoorn 2021) and we pooled the completed datasets for further analysis. We chose not to perform analysis on all five separate datasets due to the computational expense of the grid search planned, complexity of combining within and between variances for exploratory and confirmatory factor analysis, and the small percentage of tracts with missing values.

We subsampled the resulting data to provide two distinct datasets for exploratory factor analysis (EFA) and confirmatory factor analysis (CFA). Ideally, a completely new sample on the same population would be obtained to provide separate datasets for EFA and CFA (Gerbing and Hamilton 1996, Hurley et al. 1997), however, the nature of ACS data did not allow this. A distinct five-year time frame was not available to us because the number of computers variable was not included in the ACS until 2013. As a compromise, we randomly sampled states rather than random tracts for inclusion in the EFA or CFA subsample, with the assumption that spatial autocorrelation within states would provide for some differentiation between the two subsamples. The EFA subsample included 25 states and the District of Columbia (34221 census tracts) and the CFA sampled included 25 states (36845 census tracts). All data were rank transformed within sample immediately prior to factor analysis to standardize across variable units and suppress the effects of potential outliers (Yu et al. 2014, Boscoe et al. 2021). Indicator variables that were negatively correlated with deprivation (household income, gross rent, home value and education) were reverse-coded (negated) to ensure positive factor loadings.

### Exploratory Factor Analysis and Indicator Combination Reduction

We conducted EFA with one, two and three factors on the Yost indicators using the processed ACS 2014-2019 data via the `fa()` function of the `psych` package in R (Revelle 2021) to investigate potential Yost Index indicator configurations.

To find an alternative one-factor based deprivation index, we created a table containing all potential combinations of all fourteen potential indicator variables (Table 2, 14 variables, 16384 combinations). We removed combinations with less than four indicator variables to avoid non-identification by CFA later. We also limited the combinations to those that contained at least one measure of wealth (household income or poverty). We performed one-factor maximum likelihood EFA with oblique rotation on the remaining indicator combinations with the EFA states subsample data using the `fa()` function. Beta weights were determined using the regression method. We retained potential models based on a) good fit based on at least two fit indices (Tucker Lewis Index (TLI) > 0.90, fit > 0.85, corrected root mean square (CRMS) < 0.05) and RMSEA < 0.08 (Shi et al. 2019), b) balance across beta weights (all beta weights > .05 and < 0.5), and c) indicators for the factor included the three components of SES or highly correlated variables (income as household income or poverty, employment, and education as education status or computer ownership). None of the models met the assumption of multivariate normality; however, with a sample size of greater than 30,000, we relied on the central limit theorem and used fit statistics that are robust to both large sample sizes and violation of the normality assumption (Shi et al. 2019, Revelle 2021).

### Confirmatory Factor Analysis (aka Structural Equation Modeling)

Confirmatory factor analysis allows the analyst to indicate the number of factors and variables loaded on each factor as indicators; indicators are not free to load on any factor as they are with EFA. The goal of CFA is to verify the factor structure specifically defined by the analyst. We conducted CFA on factor models that emerged from the EFA model selection process using the CFA ACS 2015-2019 data subset (36845 census tracts), which we further subset into an 80/20 train/test dataset for cross-validation.

We trained the models using the 80% training dataset and the `cfa()` function in the R `lavaan` package (Rosseel et al. 2021) with arguments for maximum likelihood, standardized latent variable and standardized observations. Because the X^2^ statistic is overpowered with a sample size of just under 30,000, we also randomly sampled (without replacement) the training dataset 80 times and conducted the CFA analysis on each subsample to ensure that reliability statistics were stable on large and smaller (n ∼ 335) samples. We then checked for factor loading consistency and correlation between the final index values for train and test data models. We calculated final index values according to Boscoe et al. (2021) by grouping the factor scores into percentile values. An index value (percentile) of 100 is the least affluent, 1 is the most affluent.

We further reduced models to those with consistent train/test loadings, the comparative fit index (CFI)> 0.95, standardized root mean square residual (SRMR) < 0.08, and root mean square error of approximation (RMSEA) < 0.08 (Schermelleh-Engel et al. 2003, Shi et al. 2019). At this stage, we also checked for internal consistency (unidimensional nature) where factor loadings were all > 0.5 and all indicators included were correlated with r >|.30|) (Hair et al. 2002).

### Reliability of Final Model

Upon selection of the final model, we reaffirmed the CFA model using all processed ACS 2015-2019 census tract data (n = 71068 tracts) and relevant indicators and externally cross-checked its validity both temporally and geographically. We did this by checking loadings and fit statistics for stability between the two datasets. We also found actual index values as percentiles of factor scores for the “test” dataset and compared these to index values predicted by the externally derived model using ordinary least squares regression (R function `lm()`). For a temporal validation, we accessed and processed ACS 2011-2015 data. To check for geographic stability, we compared model performance and predicted index values between ACS 2015-2019 data for NY and MN (Berg et al. 2021).

### Validity of the Final Model

We further tested convergent validity of the model externally by relating the model index values (0-100) to various health parameters associated with affluence. We gathered these health data from the CDC PLACES census tract data-a dataset of model-based estimates human health measures at the census tract level (Centers for Disease Control et al. 2020). We found index values (percentiles of factor scores) for both our selected model ACS 2015-2019 data and found the Pearson correlation coefficient between the index values and relevant human health measures at the census tract level. Strong (r > 0.70) correlations between the Model One index and affluence related health outcomes indicated convergent validity. Specifically, we used crude prevalence estimates for self-assessed poor mental and physical health for at least two weeks, all teeth lost among adults aged >= 65, no leisure time physical activity, coronary heart disease, and diabetes. We chose these variables because they have been associated with socioeconomic status in the literature (Gilbert et al. 2003, Braveman et al. 2010, Seerig et al. 2015). We also looked at crude prevalence of basic preventative measures including basic annual checkup, “core set of clinical preventative services” for both men and women, cholesterol screening and dental visits. We repeated the above using the Yost index values for comparison.

We used R for all data processing and analysis (R Core Team 2020).

## Results

### Evaluation of the Yost Index

To determine the statistical reliability of the existing Yost index, we conducted EFA on the seven Yost indicators for the complete processed 2014-2019 ACS census tract dataset using one, two and three factors. One and two factor structures were not statistically reliable based on very poor TLI and RMSEA fit measures; however, a three-factor model was satisfactory (Tables 3, 4). Beta weights for education, employment rates and housing value/cost are low (Table 1) when constraining the variability to one factor because these indicators load more strongly on factors separate from measures of wealth (Table 4). A bivariate correlation matrix confirmed strong relationships between rent and home value, poverty, and income, working class and education and working class and income, suggesting potential redundancy in the current Yost index (Table 5).

**Table 3.**
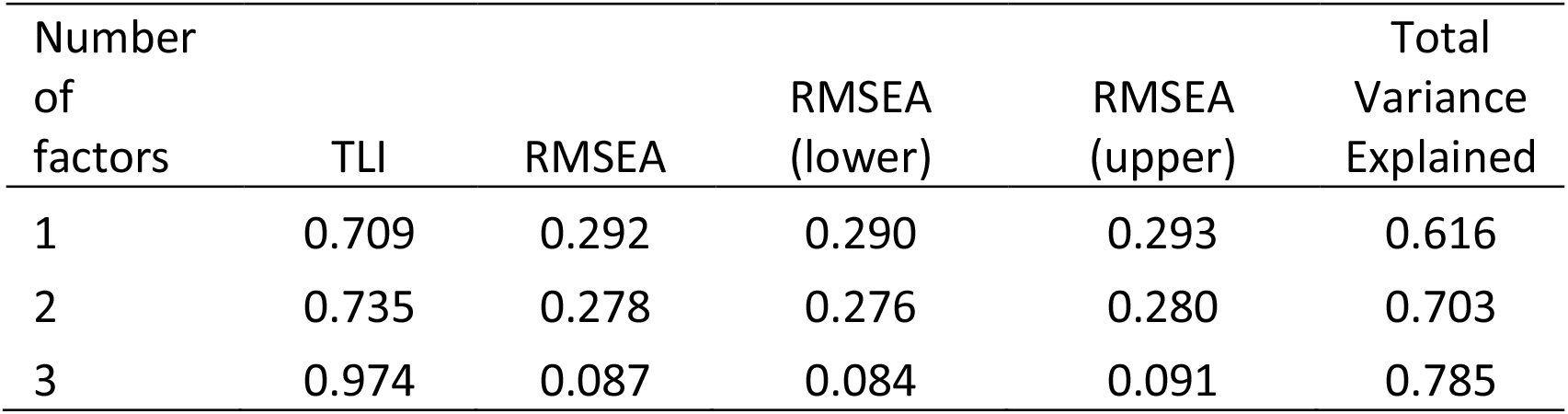
Fit measures for Yost indicators modeled as one, two and three factors using exploratory factor analysis.

**Table 4.** Factor loadings for an exploratory factor analysis three-factor structure of the seven Yost indicators. Loadings < 0.3 are disregarded.

| Indicator | Factor 1 | Factor 2 | Factor 3 |
| --- | --- | --- | --- |
| Poverty |  | .991 |  |
| Income |  | .672 | .318 |
| Home Value |  |  | .795 |
| Rent |  |  | .895 |
| Working class | 1.023 |  |  |
| Employment |  | .525 |  |
| Education | .685 |  |  |

**Table 5.** Pearson’s correlation coefficients among Yost and additional variables. Data: processed ACS 2015-2019 (without territories), N = 71066. All correlations are significant at the p < .001 level.

|  | Rent | Income | Home Value | Poverty | Working class | Employmt | Education | Gini Index | Public Asst. | Housing Cost Burden | Own Computer | Language | Health Ins. | Home Crowd |
| --- | --- | --- | --- | --- | --- | --- | --- | --- | --- | --- | --- | --- | --- | --- |
| Rent | 1.00 |  |  |  |  |  |  |  |  |  |  |  |  |  |
| Income | 0.74 | 1.00 |  |  |  |  |  |  |  |  |  |  |  |  |
| Home Value | 0.72 | 0.69 | 1.00 |  |  |  |  |  |  |  |  |  |  |  |
| Poverty | -0.48 | -0.75 | -0.40 | 1.00 |  |  |  |  |  |  |  |  |  |  |
| Working class | -0.55 | -0.74 | -0.57 | 0.64 | 1.00 |  |  |  |  |  |  |  |  |  |
| Employment | -0.23 | -0.41 | -0.22 | 0.57 | 0.38 | 1.00 |  |  |  |  |  |  |  |  |
| Education | 0.49 | 0.68 | 0.46 | -0.70 | -0.85 | -0.42 | 1.00 |  |  |  |  |  |  |  |
| GINI | -0.17 | -0.23 | 0.12 | 0.37 | -0.03 | 0.22 | -0.07 | 1.00 |  |  |  |  |  |  |
| Public Assistance | -0.15 | -0.38 | -0.18 | 0.46 | 0.44 | 0.32 | -0.48 | 0.03 | 1.00 |  |  |  |  |  |
| Housing Burden | 0.19 | -0.18 | 0.12 | 0.33 | 0.18 | 0.27 | -0.21 | 0.18 | 0.33 | 1.00 |  |  |  |  |
| No Computer | -0.61 | -0.74 | -0.50 | 0.78 | 0.71 | 0.50 | -0.80 | 0.29 | 0.37 | 0.14 | 1.00 |  |  |  |
| Language | 0.16 | -0.13 | 0.16 | 0.29 | 0.28 | 0.10 | -0.44 | 0.01 | 0.42 | 0.36 | 0.19 | 1.00 |  |  |
| Health Insurance | -0.28 | -0.46 | -0.30 | 0.52 | 0.54 | 0.23 | -0.59 | 0.07 | 0.29 | 0.12 | 0.51 | 0.47 | 1.00 |  |
| Home Crowding | 0.02 | -0.12 | 0.08 | 0.21 | 0.17 | 0.09 | -0.26 | 0.04 | 0.21 | 0.17 | 0.15 | 0.40 | 0.22 | 1.00 |

### Exploratory Factor Analysis and Indicator Combination Reduction

Our main objective was to find a statistically reliable, externally valid one-factor structure with the utility of the current Yost Index. To this end, we conducted an exhaustive investigation of indicator combinations. Indicators included were all current Yost index indicators (Table 1, 2) and proposed additional indicators (Table 2) for an initial total of 16384 indicator combinations. We reduced according to criteria outlined in the methods section resulting in a final seven combinations (Figure 1).

**Figure 1.**
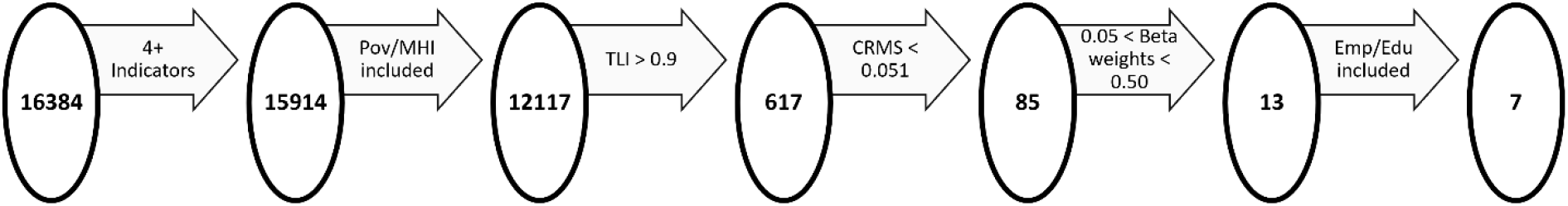
Illustration of indicator combination culling process based on inclusion of essential indicators (wealth as poverty or household income, employment status and education), fit statistics (TLI, corrected root mean square CRMS), and beta weight balance.

Seven indicator combinations emerged as potential factor models for the latent variable, deprivation (Table 6). We added an eighth model in the interest of exploration, as there were not any models with more than five indicators that met our standards and we wanted to investigate the potential of a six-indicator factor (Model 8, Table 6).

**Table 6.** Exploratory factor analysis loadings and fit parameters for the final seven (plus one) indicator combinations.

| Model | mhi | mhv | pov | emp | edu | pa | comp | hins | TLI | RMSEA | CRMS | FIT |
| --- | --- | --- | --- | --- | --- | --- | --- | --- | --- | --- | --- | --- |
| 1 |  |  | 0.49 | 0.11 | 0.35 | 0.13 |  |  | 0.98 | 0.07 | 0.03 | 0.87 |
| 2 | 0.38 |  |  | 0.09 | 0.44 |  |  | 0.17 | 0.99 | 0.04 | 0.02 | 0.88 |
| 3 |  |  | 0.48 | 0.10 | 0.33 |  |  | 0.18 | 0.97 | 0.08 | 0.04 | 0.89 |
| 4 |  | 0.09 | 0.45 | 0.09 | 0.33 |  |  | 0.16 | 0.96 | 0.09 | 0.05 | 0.86 |
| 5 | 0.31 |  |  | 0.09 | 0.47 | 0.10 |  | 0.15 | 0.96 | 0.09 | 0.05 | 0.86 |
| 6 |  |  | 0.44 | 0.09 | 0.32 | 0.11 |  | 0.16 | 0.96 | 0.09 | 0.05 | 0.88 |
| 7 |  |  | 0.26 | 0.05 | 0.31 |  | 0.36 | 0.10 | 0.92 | 0.15 | 0.05 | 0.92 |
| 8 |  |  | 0.27 | 0.06 | 0.31 | 0.06 | 0.31 | 0.10 | 0.90 | 0.15 | 0.06 | 0.90 |

Abbreviations: mhi: median household income, mhv: median home value, pov: % earning < 150% poverty rate, emp: % older than 16 unemployed, edu: education level, pa: receive public assistance income (%), comp: no laptop or desktop computer (%), hins: no health insurance (%)

### Confirmatory Factor Analysis

We conducted CFA on these eight indicator combinations. We constrained all models to one factor and tested them using the entire training dataset (80% of the CFA subset, n ≈ 29476) as a whole and as 80 random subsamples. Means of the subsample fit parameters and factor loadings did not vary considerably-all models were stable across runs. Model One emerged as the best fit model while models three, five and six had nearly satisfactory fit (Table 6). However, internal consistency/unidimensional character of a factor requires that item to item correlation is greater than r = 0.30 (Hair et al. 2002), which excluded models three, five and six, because health insurance (hins) is poorly correlated with employment (r= 0.23) and public assistance (r= 0.29, Table 5). Thus, we evaluated only Model One. Cross-validation between the 80% training dataset and 20% test dataset resulted in excellent agreement, with the main differences in parameters being a result of sample size (Table 7).

**Table 7.** Model One factor loadings and fit statistics for train/test (80/20%) cross-validation datasets.

|  | Train | Test |
| --- | --- | --- |
| pov | 0.92 | 0.91 |
| emp | 0.56 | 0.57 |
| edu | 0.80 | 0.80 |
| pa | 0.61 | 0.61 |
| CFI | 0.99 | 0.99 |
| SRMR | 0.02 | 0.02 |
| RMSEA | 0.06 | 0.06 |
| RMSEA (upper) | 0.07 | 0.08 |
| RMSEA (lower) | 0.06 | 0.05 |
| Absolute Deviations (index) | 0.28 |  |
| Index Prediction R2 | 0.9996 |  |

Abbreviations: pov: % earning < 150% poverty rate, emp: % older than 16 unemployed, edu: education level, pa: receive public assistance income (%)

### Reliability of Final Model

We also cross-validated Model One using external temporally and geographically distinct datasets. To verify reliability over time we used ACS 2011-2015 data, processed in the same way as the original ACS 2015-2019 data. We based assessment of temporal consistency using factor loading and fit parameter stability, as well as the relationship between percentiles of actual factor scores and predicted factor scores (index values, Table 8). Loadings for employment and public assistance differed between the time frames and the fit was not as strong for Model One when run on 2011-2015 data; however, the fit was still good, all loadings are still > 0.5, and index values for the census tracts were very consistent.

**Table 8.** Comparison of Model One loadings, fit statistics, and index values between two temporally distinct datasets (ACS 2011-2015, 2015-2019) and two spatially distinct datasets (MN, NY from ACS 2015-2019).

|  | 2015-2019 | 2011-2015 | MN | NY |
| --- | --- | --- | --- | --- |
| pov | 0.92 | 0.91 | 0.97 | 0.88 |
| emp | 0.57 | 0.66 | 0.51 | 0.55 |
| edu | 0.80 | 0.81 | 0.67 | 0.84 |
| pa | 0.61 | 0.58 | 0.47 | 0.71 |
| CFI | 1.00 | 0.99 | 0.96 | 1.00 |
| RMSEA | 0.06 | 0.08 | 0.15 | 0.05 |
| RMSEA (upper) | 0.07 | 0.08 | 0.19 | 0.07 |
| RMSEA (lower) | 0.06 | 0.07 | 0.12 | 0.03 |
| Mean Absolute Deviation | 0.77 |  | 6.01 | 5.89 |
| Actual vs Predicted Index R<sup>2</sup> | 0.9986 |  | 0.92 | 0.93 |

Abbreviations: pov: % earning < 150% poverty rate, emp: % older than 16 unemployed, edu: education level, pa: receive public assistance income (%)

To check for geographic consistency of Model One, we found Model One on subsets of the 2015-2019 ACS data consisting of NY and MN census tracts and looked for reliability of loadings, parameters and factor score prediction (Table 8). Model One is less convincing on MN data and factor loadings differ, although the unidimensional status is retained for both states. Index prediction between the two states is still quite good; because the index is based on percentiles, it is likely robust to minor discrepancies in fit.

### Validity of the Final Model

To further check convergent external validity of Model One, we wanted to associate the index to a completely different, but related dataset (Hair et al. 2002). We included the Yost Index value associations for comparison. Using the modeled tract level human health data from the CDC PLACES dataset (Centers for Disease Control et al. 2020), we were able to relate Model One and Yost index values to public health measures at the tract level (Table 9). Relationships between Model One and mental health, physical health, teeth loss, and access to leisure time physical activity were quite strong; however, the relationships were usually slightly stronger with the Yost Index. Preventative measures, which tend to be less accessible to those of lower SES (Singu et al. 2020) were more strongly related to Model One; however, relationships between these health measures and our index and the Yost Index were a bit weak, apart from dental visits (Table 9).

**Table 9.** Correlation (Pearson’s r) between index values for Model One and the Yost index and various PLACES data human health outcomes/measures.

| Model | One | Yost |
| --- | --- | --- |
| Mental Health | 0.85 | 0.85 |
| Physical Health | 0.83 | 0.85 |
| Tooth Loss | 0.86 | 0.87 |
| Coronary Heart Disease | 0.50 | 0.61 |
| Diabetes | 0.72 | 0.74 |
| Leisure Physical Activity | 0.84 | 0.85 |
| Checkup | -0.05 | 0.03 |
| Core Men's Preventative Health | -0.69 | -0.61 |
| Core Women's Preventative Health | -0.67 | -0.63 |
| Cholesterol Screening | -0.60 | -0.59 |
| Dental Visit | -0.90 | -0.88 |

Finally, we compared the Model One and Yost index values (ACS 2015-2019) using ols regression and investigated outliers to better understand how the models differ. We considered a difference in index values of greater than 30 percentiles to be “poor agreement” (Boscoe et al. 2021). The difference between Model One and Yost index values exceeded 30 percentiles in 500 census tracts (0.7%), 70.2% of which were tracts in California or New York. We randomly selected ten of these census tracts to investigate (Table 10). In general, census tracts estimated to be less affluent by the Yost index but a higher level of affluence by the Model One index were associated with areas with low cost of housing and/or low income, but with low poverty and/or low public assistance use, such as retirement communities. Census tracts with a more affluent Yost index than Model One index value were commonly located in primarily working-class neighborhoods with very high cost of living/home value/rent and high public assistance.

**Table 10.** Characteristics of a sample of census tracts with poor agreement between Model One and Yost index values.

| Census Tract | Yost Index -<br>Model One<br>Index | Characteristics |
| --- | --- | --- |
| Census Tract 1993, Los Angeles County, California | -47 | Income high, home values very high, public assistance very high |
| Census Tract 5306.02, Los Angeles County, California | -45 | Home values, rent and income high, mostly working class, high unemployment, low education, very high public assistance |
| Census Tract 47.17, Ventura County, California | -40 | Home values, rent, income high, low education, high poverty, high proportion working class |
| Census Tract 1459.02, Suffolk County, New York | -37 | Working class neighborhood; Income high, rent very high, home value high, education low, public assistance and working class very high |
| Census Tract 1002, Shawano County, Wisconsin | 32 | Income, rent low, unemployment very low, home value medium low, poverty, public assistance low |
| Census Tract 128.03, Polk County, Florida | 34 | Retirement community: Home value, rent, income low, unemployment and public assistance also low |
| Census Tract 44.04, Pima County, Arizona | 38 | Retirement community; home values and income low, but public assistance also very low |
| Census Tract 26.06, Marion County, Florida | 40 | Retirement community; Home value and income low, unemployment, public assistance low, education high |
| Census Tract 506.04, Indian River County, Florida | 43 | Home values, rent and income very low, but poverty is ranked in the middle, public assistance very low |
| Census Tract 9653, Windsor County, Vermont | 44 | Manufacturing town: Low rent, income, home value ranked in the middle, education very high, public assistance low, poverty ranked middle |

## Discussion

Changing technologies and economics require re-evaluation and potential revision of SES and other indices over time. Such index updates should be based on reliable and reproducible methods such as empirical analysis or “subjective” methods. Of the many methods one could use, we chose to include many ACS derived potential indicators that are related to SES in a grid search to find the best-fit single-factor structure to describe the latent variable deprivation/SES/affluence. We limited ourselves to one factor in the interest of simplicity, parsimony, ease of replication, and utility for the public health communities. The Model One Index we developed is a spatially and temporally reliable index based on a statistically valid factor with poverty, education, public assistance, and unemployment as indicators.

Both the Model One and Yost Indices correlate well with at least some health measures associated with affluence including tooth loss, self-assessed mental and physical health, access to leisure physical activity and diabetes.

As it is, the Yost index is a valuable covariate that improves model fit or helps to explain relationships between SES and healthcare and/or outcomes (Butler et al. 2020, Li et al. 2021, Liu et al. 2021); however, the Yost Index is based on a statistically unreliable one-factor structure with unbalanced weights. Inclusion of low weighted indicators (e.g., working class, unemployment, education) gives the impression that those indicators do little to predict the latent variable; however, if all relevant factors are considered, they are influential. To include the explanatory power of working class, unemployment and other low weighted indicators, the Yost Index should be based on a three-dimensional structure.

Thus, while the Yost Index, as a black box model, has value, it is more valid to call the indicator weights subjective-based on expert opinion, than to call them factor analysis beta weights. Despite its statistical faults, the Yost Index seems to be well associated with deprivation and is also well correlated with SES related human health measures.

The index output of our proposed model (Model One) and the Yost Index are quite similar. When comparing poor agreement index values for tracts, the main differences are a function of housing cost, and possibly cost of living as a result. Working class neighborhoods in LA County, for example, have a higher affluence rating than they would on the Model One index due to the extremely high housing costs in that portion of California. Retirement communities in FL and AZ exemplify tracts where the Model One index indicates higher affluence than the Yost index; citizens may be living on fixed incomes, but they are differentiated from actual impoverished locations by the low poverty levels and low public assistance. The Yost index is sensitive to regional housing costs because it employs two housing cost variables, both of which are likely to influence local household incomes. The Model One index is likely sensitive to local politics due to its reliance on ACS survey participant disclosure of public assistance.

While public assistance in the index seems to control for cost-of-living disparities, if participants do not request/disclose public assistance, the results will be skewed. Both indices may be better on a regional scale since home values and public opinion are generally spatially autocorrelated.

While they are similar, overall, the Model One Index has advantages over the Yost Index on a large geographical scale. The Model One Index is more robust to extremes in local cost of living conditions, is comprised of ACS variables that rarely, if ever, require imputation by the end-user, and is a more parsimonious solution with a true one-factor structure. However, there are many deprivation indices in the literature; the choice of index will likely depend on the researcher’s definition of socioeconomic status and deprivation.

### Limitations and Future Work

By forcing a one factor structure, we necessitated correlation among the indicators included in the final model. Although the simplicity of the final index enhances utility, this approach may have limited the potential descriptive ability of the index; many potential indicators of deprivation are not strongly correlated with wealth, employment, or education. Using structural equation modeling to allow for more than one factor may provide a more descriptive index of deprivation.

Rank transformation of the ACS data prior to scaling and modeling results in a loss of information, but a gain in outlier suppression. Rank transformed data do not correlate as well over time as do raw data, and we may have lost some temporal transferability by ranking the raw data while developing Model One. While Model One does seem to perform well over time, its performance may be even better if using raw data. An investigation into the effects of rank transformation would benefit the index and development of such indices in the future.

The US Census ACS data are not perfect and have inherent variability. We did not incorporate this potential source of uncertainty in our models. Boscoe et al. (in press) calculated confidence intervals around the Yost Index values using ACS variance replicate tables and reported that the index at the percentile level may be misleading due to uncertainty around the Yost Index values. The average tract index estimate precision was +/-eight percentiles, but some tracts, especially tracts near the median, had much larger confidence intervals. Future work on Model One should incorporate uncertainty in the data.

## Data Availability

All data used in the study are available publicly via the US Census Bureau, IPUMS NHGIS and/or CDC.

https://www.cdc.gov/places/index.html

https://www.nhgis.org/

https://www.census.gov/programs-surveys/acs/

## Acknowledgements

We thank Drs. Valerie Haley, Darren Gemoets, and Thomas O’Grady for editorial comments on an earlier version of this paper.

